# Is the Sars-CoV-2 virus a possible trigger agent for the development of achalasia?

**DOI:** 10.1101/2022.09.19.22280068

**Authors:** Janette Furuzawa-Carballeda, María Eugenia Icaza-Chávez, Diana Aguilar-León, Norma Uribe-Uribe, María del Carmen Nuñez-Pompa, Alonso Trigos-Díaz, Rodrigo Areán-Sanz, Dheni A. Fernández-Camargo, Coss-Adame E Enrique, Miguel A. Valdovinos, Eduardo Briceño-Souza, Luis A. Chi-Cervera, Miriam Olivares-Flores, Gonzalo Torres-Villalobos

## Abstract

**BACKGROUND:** Previous studies have suggested that achalasia is an autoimmune disease whose probable causal agent is a neurotropic virus that chronically infects the myenteric plexus of the esophagus and, in a genetically susceptible host, induces the disease. The association between achalasia and coronaviruses has not been reported in the literature.

**AIMS:** To evaluate the presence of the SARS-CoV-2 virus, the ACE2 expression, the tissue architecture, and immune response in the lower esophageal sphincter muscle (LESm) of achalasia patients who had SARS-CoV-2 (achalasia-COVID-19) infection before laparoscopic Heller myotomy (LHM) and compare the findings with type II achalasia patients and transplant donors (controls) without COVID-19.

**METHODS:** The LESm of 7 achalasia-COVID-19 patients (diagnosed by PCR), ten achalasia patients, and ten controls without COVID-19 were included. The presence of the virus was evaluated by *in situ PCR* and immunohistochemistry. ACE2 receptor expression and effector CD4 T cell and regulatory subsets were determined by immunohistochemistry.

**RESULTS:** Coronavirus was detected in 6/7 patients-COVID-19. The SARS-CoV-2 was undetectable in the LESm of the achalasia patients and controls. The ACE2 receptor was expressed in all the patients and controls. One patient developed achalasia type II post-COVID-19. The percentage of Th22/Th17/Th1/pDCreg was higher in achalasia and achalasia-COVID-19 pre-HLM vs. controls. The Th2/Treg/Breg cell percentages were higher only in achalasia vs. controls.

**CONCLUSION:** The presence of the SARS-CoV2 and its receptor in the LESm of type II achalasia-COVID-19 patients but not in the controls suggests that it could affect the myenteric plexus. Unlike achalasia, patients-COVID-19 have an imbalance between effector CD4 T cells and the regulatory mechanisms.

---

Idiopathic achalasia is a motor disorders of the esophagus. It has a prevalence of 8 cases per million inhabitants and an estimated incidence of 0.3-1.63 per 100,000/year.^1^ Achalasia is characterized by aperistalsis, insufficient relaxation of the lower esophageal sphincter (LES) in response to swallowing, and an increase in the resting tone of the sphincter.^2^ It is typically characterized by dysphagia for solids and liquids. Other clinical manifestations, such as regurgitation, retrosternal pain, cough, aspiration, weight loss, and nutritional deficiencies, may coexist.^3^ This motor disorder results from the loss of neurons of the myenteric esophageal plexus due to inflammatory destruction. The precise causes of this condition have been explored and are still unclear.^4^ Factors, including infections, autoimmunity, and genetic components, can contribute to the development of the inflammatory process. Moreover, the high prevalence of specific antibodies against the myenteric plexus, the increased serum pro-inflammatory cytokines levels, and chronic inflammatory infiltrates in the LES of achalasia patients support its association with autoimmune disease.^5,6^ The geographic profile may reflect a genetic predisposition. It has been determined that genes of the major histocompatibility complex class II (HLA-DRB1 * 14: 54 and DQB1 * 05: 03) and the conserved haplotype DRB1 * 14: 54-DQB1 * 05: 03 confer risk for the development of achalasia in mestizo Mexicans. It could be important for developing an aberrant immune response secondary to initiates such as viral or bacterial infections.^7^ Viral infections, especially chronic latent and active infections caused by neurotropic viruses with a preference for squamous epithelium, could be a triggering factor in the chronic inflammatory process of the myenteric plexus of the esophagus in genetically predisposed subjects.^6,8^ Proposed candidate viruses have included herpes simplex virus (HSV), varicella Zoster virus, Measles virus, human papillomavirus, “JC” virus, mumps, and bornavirus. The association of achalasia with other viruses such as Cytomegalovirus, Epstein-Barr virus,^6,8,^ or Coronaviruses has not been reported in the literature. However, there is a high susceptibility to infection of the gastrointestinal tract by SARS-CoV-2 due to the expression of the receptor for angiotensin-converting enzyme type II (ACE2) in ileal epithelial cells (∼30% ACE2-positive cells) and esophagus epithelial cells (>1% ACE2-positive cells).^9^ Moreover, SARS-CoV-2 RNA has been detected in mucosal biopsies of the esophagus in COVID-19 patients.^10^

Thus, this study was focused on the evaluation of the presence of the SARS-CoV-2 virus, the ACE2 expression, the tissue architecture, and immune response in the LES muscle of patients with achalasia who had COVID-19, confirmed by PCR, at least one month before surgery and compare the findings with patients with type II achalasia and transplant donors (controls) from the pre-COVID-19 period.

## Methods

### Patients

This was a cross-sectional, observational study comparing LES muscle biopsies from type II achalasia patients (n=7) who had COVID-19 at least one month before LHM (2020-2022), with type II achalasia patients (n=10) and with transplant donors (n=10) from the pre-COVID period (2015-2018). Tissue samples were used for histology, *in situ* PCR, and immunohistochemistry. All patients were recruited from the Outpatient Clinics of Gastroenterology and Surgery of the Instituto Nacional de Ciencias Médicas y Nutrición Salvador Zubirán (a tertiary referral center in Mexico City, Mexico). Esophagram, HRM (classified based on Chicago v4.0), and endoscopy were performed to diagnose achalasia.^11,12^ Patients were excluded from the study if they presented any of the following diagnoses: Chagas disease, eosinophilic esophagitis, esophageal stricture, gastric or esophageal cancer, scleroderma, hiatal hernias, gastroesophageal reflux disease with erosive esophagitis, human immunodeficiency virus (HIV), or hepatitis C virus (HCV) infections.

None of the patients had been taking opioids. None of the transplant donors had previously known metabolic, inflammatory, neoplastic, or autoimmune diseases. No viral infections with cytomegalovirus (CMV), hepatitis C virus (HCV), hepatitis B virus (HBV), and human immunodeficiency virus (HIV) were detected. Tests for syphilis and serum ANAs were negative. The cause of death in 9 of the donors was brain injury, three by subarachnoid bleeding, and four by hemorrhagic stroke.

Demographic, clinical, and laboratory information was collected (Table 1).

**Table 1.**
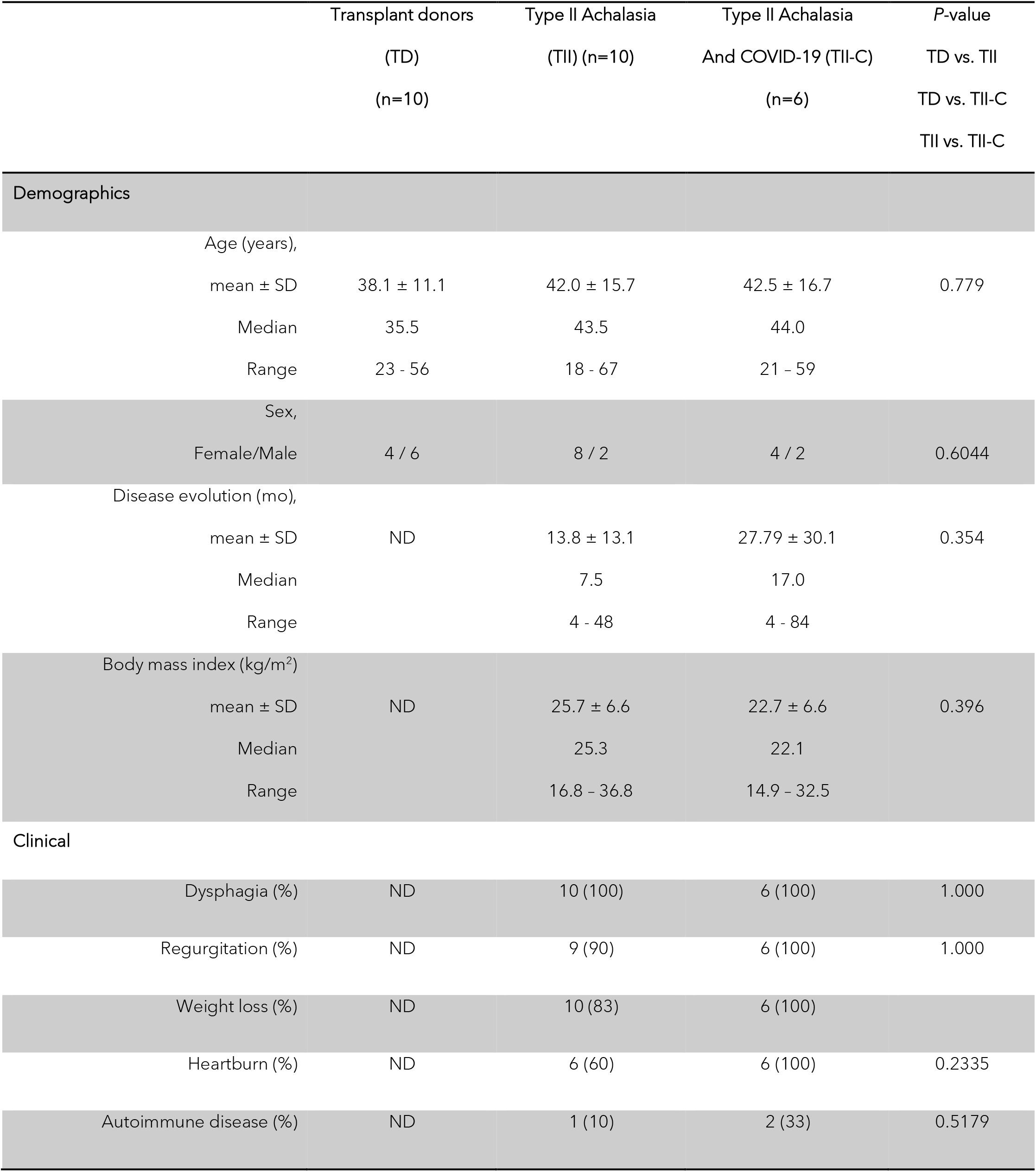
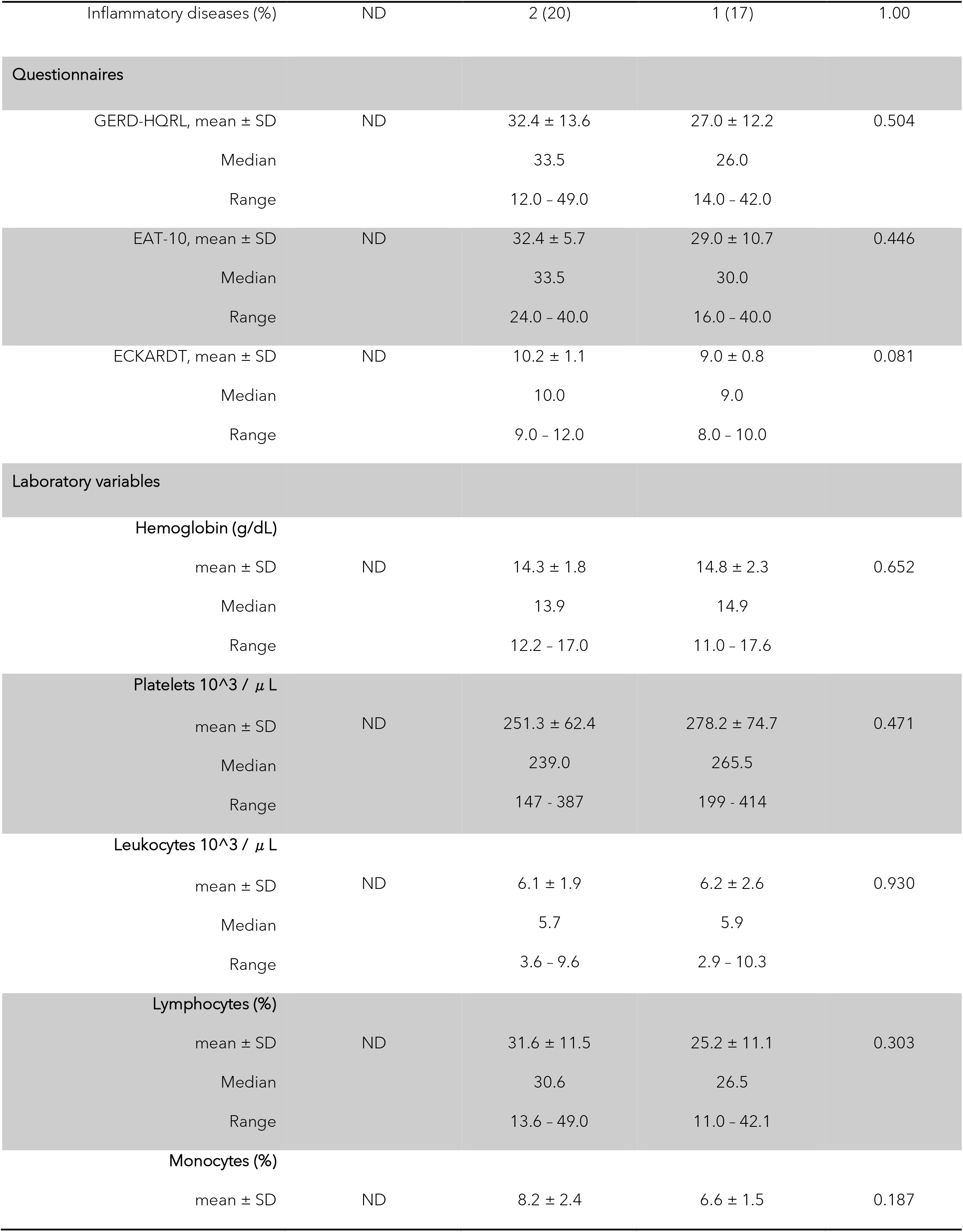
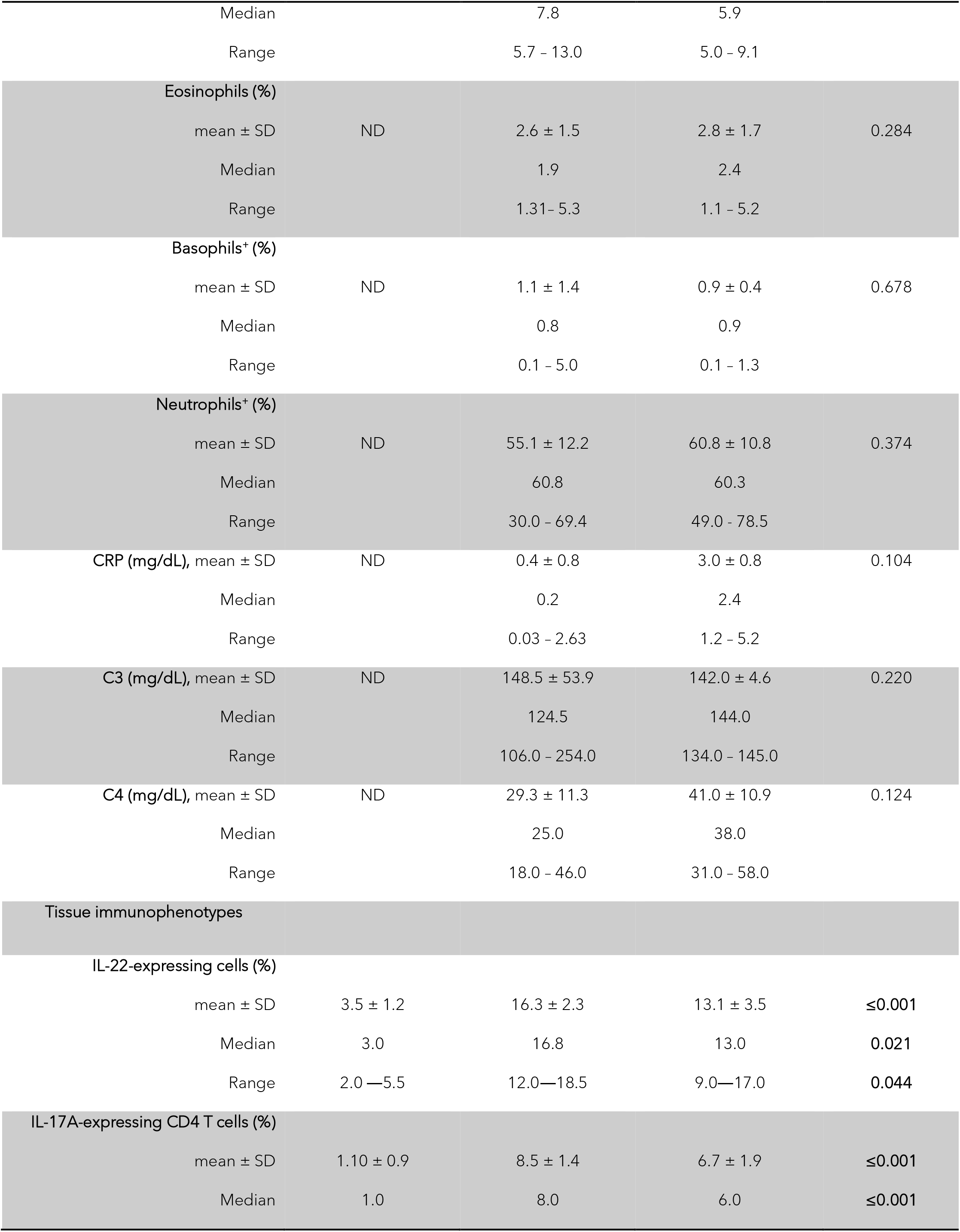
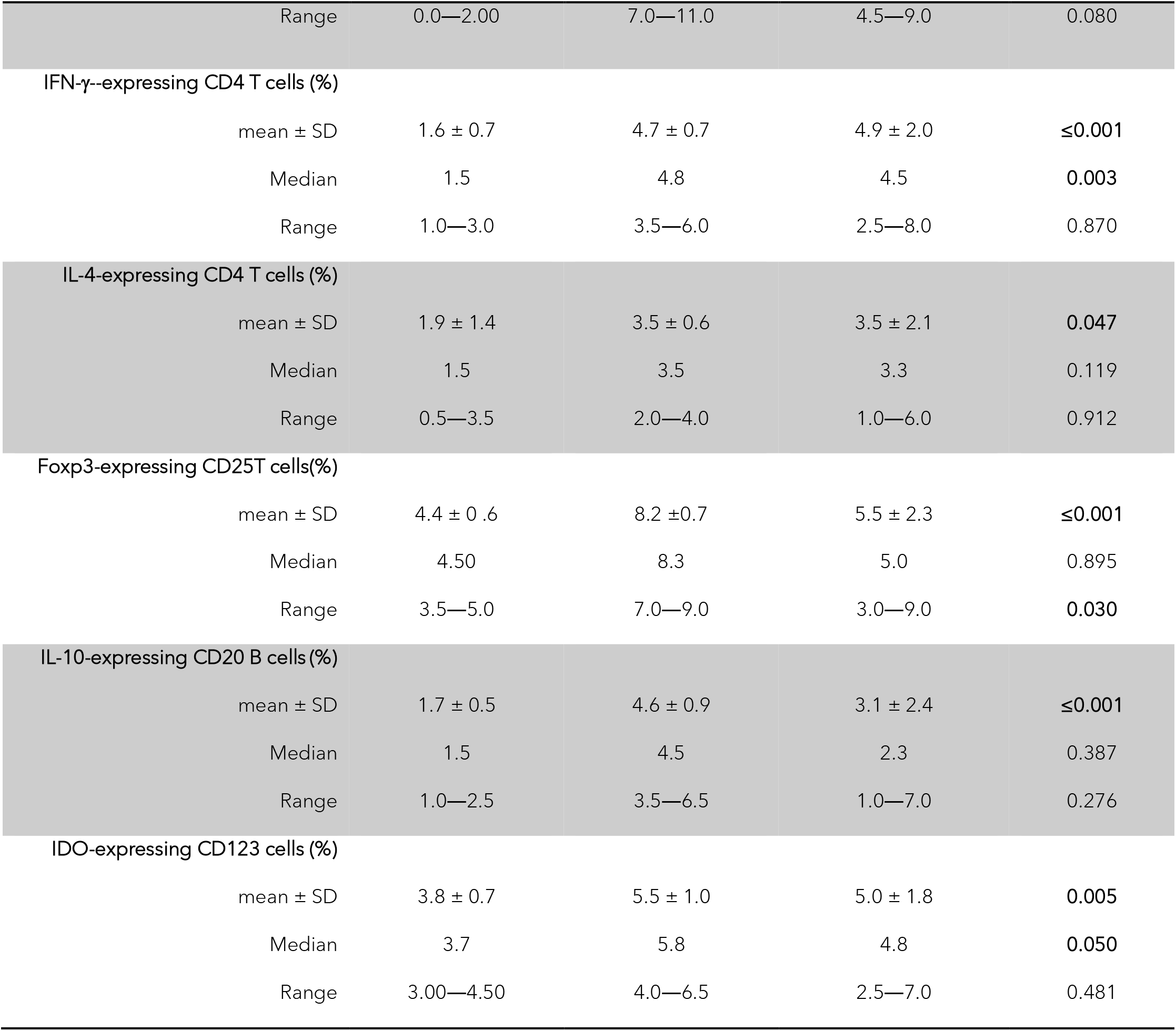
Demographic, clinical, and laboratory variables

### Esophageal High-Resolution Manometry

An esophageal HRM was performed in the achalasia patients before being referred for surgery. A solid-state HRM probe with 36 circumferential sensors was used (Medtronic©, Minneapolis, MN, USA). Having the patient in a sitting position and at 45 degrees, stationary esophageal HRM was performed. After a 12-hr. fasting period, the probe was inserted trans-nasally until passing the esophagogastric junction and assessed visually on the computer screen. Ten water swallows of 5 mL separated by 30 seconds were provided. Analyses were performed using Manoview 2.0 (Medtronic©). Patients were classified according to the latest Chicago classification v4.0^11,12^ into four groups: (a) type I achalasia (without pressurization within the esophageal body), (b) type II (with >20% panpressurization), and (c) type III (spastic). Two gastroenterologists (EC-A, MAV) experts in esophageal HRM performed the classification.

### HRM Data Analysis

Analysis was performed with ManoView software (Given Imaging, Yokneam, Israel). Four critical metrics of HRM, the distal contractile integral (DCI), the basal EGJ pressure, the intrabolus pressure (IBP), and the integrated relaxation pressure (IRP), were used to assess the pressure motor activity of the esophageal body and EGJ. Achalasia was defined by IRP >15 mm Hg and aperistalsis. Type I achalasia was defined by absent peristalsis with no compartmentalization of intrabolus pressure. Type II achalasia was defined as panesophageal pressurization in at least 20% of swallows in a 30-mmHg isobaric contour. Type III achalasia was characterized by premature contractions with shortened distal latency (<4.5 sec) in at least 20% of swallows.

### Tissue biopsies

Biopsies of the LES muscle of achalasia and transplant donors were taken during LHM. After the myotomy was completed without using energy devices, a full-thickness muscle biopsy (2 mm wide and 2 cm long) was obtained by cutting with scissors and was immediately preserved in formalin. The LES of transplant donors was included as tissue control. The EGJ was obtained during organ procuration, previously signed informed consent from the family. The EGJ with 3 cm of the esophagus and 2 cm of the stomach was taken. The tissue was transported at 4°C in Bretschneider’s (Custodiol) solution for 4–6 h. Subsequently, a full-thickness biopsy of the esophagus muscle (including the myenteric plexus) was obtained. Tissue was immediately formalin-fixed and paraffin-embedded.

### Immunohistochemistry

Four *μ*m thick tissue sections were deparaffinized and rehydrated. Then enzymatic antigen retrieval was performed. Tissues were incubated for one h at room temperature with mouse monoclonal anti-SARS-CoV/SARS-Cov2 (COVID-19) spike antibody (GeneTex, Inc., USA, CA), goat polyclonal anti-human IL-22 antibody, mouse monoclonal IgG_1_ herpes simplex virus-1 (HSV-1) antibody (Santa Cruz Biotechnology, Santa Cruz, CA, USA) and ACE2 (Bio-SB, Santa Barbara CA, USA) at ten *μ*g/mL. Binding was detected by incubating sections with biotinylated donkey anti-goat IgG antibody (DAKO LSAB+System-HRP Glostrup, Denmark). Slides were incubated with 3,3’-diaminobenzidine (DAB) (Sigma-Aldrich) and counterstained with hematoxylin. Negative control staining was performed with normal human serum diluted at 1:100 instead of primary antibody. The IHC universal negative control reagent design to work with rabbit, mouse, and goat antibodies (IHC universal negative control reagent, Enzo Life Sciences, Inc., Farmingdale, NY, USA) was also used as a negative control. The reactive blank was incubated with phosphate buffer three saline-egg albumin (Sigma-Aldrich) instead of the primary antibody. Controls excluded non-specific staining or endogenous enzymatic activities. For each biopsy, at least two different sections and two fields (×320) were examined. IL-22 immunostained cells were reported as the percentage of immunoreactive cells. Results are expressed as the mean ± standard error of the mean (SEM).

### Double-Staining Procedure

To determine the subpopulation of CD4^+^/IL-17A^+^ (Th17), CD4^+^/IL-4^+^ (Th2), CD4^+^/IFN-*γ*^+^ (Th1), CD25^+^/Foxp3^+^ regulatory T cells (Tregs), CD20^+^/IL-10^+^ B cells (Bregs), and CD123^+^/IDO^+^ plasmacytoid dendritic cells (pDCregs) a simultaneous detection was performed (MultiView (mouse-HRP/rabbit-AP) Enzo Life Sciences). Before staining, sections were deparaffinized with xylene and rehydrated with a graded series of ethanol and water washes. Following standard dewaxing and rehydration, enzyme antigen retrieval was performed (Enzo Life Sciences, Inc., Farmingdale, NY, USA). Tissues were blocked with a peroxidase solution. Then non-specific background staining was avoided with the IHC background blocker (Enzo Life Sciences). Samples were incubated with rabbit polyclonal anti-CD4, anti-CD20, anti-CD25 IgG (Santa Cruz Biotechnology), anti-CD123 IgG antibody (Abcam pcl, CA, UK), or mouse monoclonal anti-IL-17A, anti-IL-4, anti-IFN-*γ*, anti-IL-10, anti-Foxp3 IgG1, or anti-IDO IgG antibody (Santa Cruz Biotechnology) at ten *μ*g/mL during 30 min at room temperature. Slides were washed and incubated with PolyView IHC reagent (mouse-HRP) and PolyView IHC reagent (rabbit-AP) for 20 min. Finally, antigens were visualized using HRP/DAB and alkaline phosphatase (AP)/Permanent Red. Tissues were counterstained with hematoxylin and mounted in an aqueous mounting medium. Double positive cytokine-expressing cells (burgundy cells) were assessed in three fields (×320) and were reported as the percentage of immunoreactive cells. Results are expressed as the mean ± standard error of cells’ mean (SEM). Negative controls staining and reactive blank were performed as described previously.^6^

### In situ RT-PCR

Four-µm-thick sections of available formalin-fixed paraffin-embedded tissue were deparaffinized and rehydrated. Cells were permeabilized and incubated with one mg/L proteinase K (Promega, Madison, WI, USA) for 30 min at 37°C. Tissue was treated with DNase I (Invitrogen, Alameda, CA, USA). RT-PCR was done by incubation of the sections with 1 mM Magnesium sulfate, 1 ml SuperScript II RT/Platinum Taq DNA polymerase mix (Invitrogen, Alameda Ca, USA), dNTPs 0·2 mmol/L dUTP-digoxigenin (Boehringer Mannheim, Lewes, UK), and 10 mM of each primer E-F(ACAGGTACGTTAATAGTTAATAGCGT), E-R (ATATTGCAGCAGTACGCACACA), Rd-R(GTGARATGGTCATGTGTGGCGG), and Rd-F (CARATGTTAAASACACTATTAGCATA). The slides were sealed with the Assembly tool (Perkin Elmer) and placed in a TouchDown thermocycler (Perkin Elmer, Cambridge, UK), with the following conditions of RT-PCR: 1 cycle at 55°C for 15 min, 1 cycle at 95°C during 3 min, 40 cycles at 95°C for 15 seg, and 1 cycle at 58°C during 60 seg, and finally at 12°C. PCR products were detected with alkaline phosphatase-conjugated sheep antibodies against anti-digoxigenin (Roche Mannheim Germany) diluted 1/500, and the chromogen was 5-bromo-4-chloro-3-3 indolyl phosphate toluidine salt, and tetrazolium nitroblue (Boehringer Manheim) diluted 1/50. Sections were counterstained with nuclear fast red or malachite green to avoid interference with the blue signal generated by SARS-CoV2 DNA. One section from one block from each individual was used in masked assays to avoid bias, and it was scored positive if the blue reaction product was seen. One section from a patient with SARS-CoV2 infection was used as a positive control. The negative controls included ten biopsies of type II achalasia patients and ten biopsies of transplant donors obtained before the COVID-19 period (2015-2018).

### Statistical analysis

Descriptive statistic was performed, and categorical variables were compared using the *χ*2 test or Fisher’s exact test. Immunohistochemical statistical analysis was performed using a one-way analysis of variance on ranks by the Holm-Sidak method or Dunn’s test for all pairwise multiple comparison procedures. Statistical analyses were performed using the Sigma Stat 14.5 program (Aspire Software International, Leesburg, VA, USA). Data are expressed as the median, range, and mean ± standard deviation (SD)/ standard error of the mean (SEM). *P* values less than or equal to 0.05 were considered significant.

### Ethical Considerations

This work was performed according to the principles expressed in the Declaration of Helsinki. The ethical committee from the Instituto Nacional de Ciencias Médicas y Nutrición Salvador Zubirán approved the study, and written informed consent was obtained from all subjects (Ref. No. 1522).

## Results

### Patient characteristics

Age, sex, disease duration, laboratory data of patients with achalasia, patients with achalasia who had COVID-19, and healthy controls are shown in Table 1. There were no statistically significant differences between groups

The clinical manifestations in patients with achalasia and the prevalence of autoimmune or inflammatory disease comorbidity were similar (Table 1).

Six of seven patients with achalasia who had COVID-19 pre-LHM (diagnosed with a positive result of real-time reverse-transcription polymerase chain reaction for SARS-CoV-2) had the virus in the LES muscle (Fig. 1D and E). Patients had suggestive symptoms such as fever, headache, cough, or dyspnea, plus at least other symptoms such as malaise, myalgias, arthralgias, rhinorrhea, throat pain, conjunctivitis, vomiting, or diarrhea. Five of six adult outpatients had mild to moderate symptomatic COVID-19. They did not receive supplemental oxygen via nasal cannula or invasive mechanical ventilation.

**Figure 1.**
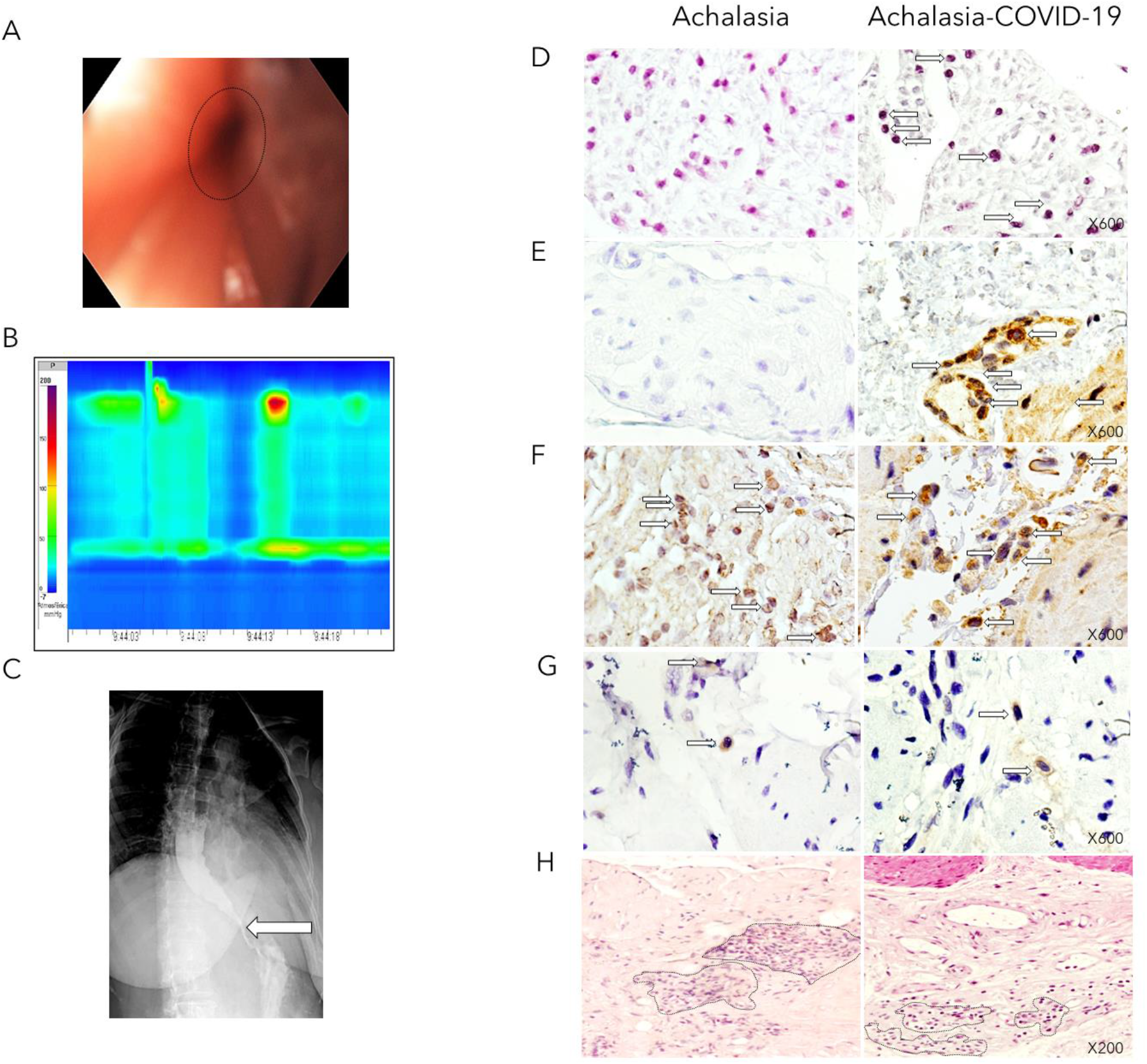
(A) Upper endoscopy shows dilated esophagus (patient that developed achalasia post-COVID-19). (B) High-resolution manometry shows abnormal relaxation of the lower esophageal sphincter (LES) and aperistalsis with panesophageal pressurization in 40% of swallows, corresponding with type II achalasia. (C) Barium esophagram that shows dilation and bird beak sign (arrow). (D) The photographs depict the *in situ* PCR for SARS-CoV2 (arrows indicate positive cells in dark blue) of the LES muscle of type II achalasia patient (*left*) and type II achalasia post-COVID-19 patient (*right*). Magnification x600. (E) The left and right panels show immunohistochemistry for SARS-CoV2 (arrows indicate positive spike expression in brown). Magnification x600. (F) The left and right panels show immunohistochemistry for ACE2 expression (Arrows depict positive cells in brown). Magnification x600. (G) The left and right panels show immunohistochemistry for HSV-1 expression (Arrows depict positive cells in brown). Magnification x600. (H) Tissue architecture of the LES muscle of type II achalasia patient (*left*) and type II achalasia post-COVID-19 (*right*). Dot circles depict inflammatory infiltrates. Magnification x200.

Only one patient was hospitalized. The patient had a history of obesity, atopic dermatitis, allergic rhinitis since childhood, and asthma. The patient was diagnosed with SARS-CoV-2 pneumonia by PCR and hospitalized. The patient had dyspnea, vomiting, nausea, headache, malaise, arthralgias, dry cough, chest pain, chills, and fever. The CT scan showed multiple ground-glass opacities with a peripheral and bronchovascular distribution, affecting between 25 to 50% of the lungs.

The patient was treated with low molecular weight heparin, dexamethasone 6 mg/24 hours, 4th generation cephalosporin, IV omeprazole, inhaled steroid LABA, and low-flow supplemental oxygen via nasal cannula. After 72 h, the patient was discharged and treated with linezolid, bromhexine/oxeladine, paracetamol, dexlansoprazole, alloglutamol, sucral, intranasal budesonide/formoterol. Three months later, patient had dysphagia to solids with cough, regurgitation, heartburn, and vomiting. The HRM showed an IRP of 15.4 mmHg at 4 seconds with panesophageal pressurization in 90% of all swallows, confirming the diagnosis of type II achalasia (Fig. 1A, B, and C).

### HSV-1, SARS-CoV2 and ACE2 receptor identification

SARS-CoV2 identification was performed by *in situ* PCR and immunohistochemistry. Only the patients with achalasia who had COVID-19 before LHM, but neither achalasia nor transplant donors showed SARS-CoV2 infected mononuclear and endothelial cells (Fig. 1 D and E). The ACE2 receptor was expressed in all tissues of transplant donors and patients with achalasia regardless of whether they had been previously infected with the SARS-CoV-2 virus (Fig. 1F). The HSV-1 virus was detected in all tissues of achalasia and achalasia-COVID-19 patients (Fig. 1G)

### Histological findings

The histological characteristics of the LES muscle of the patients with type II achalasia who had COVID-19 before LHM, biopsies from patients with type II achalasia, and transplant donors from the pre-COVID-19 period were performed (Fig. 1 H). The histological findings showed that transplant organ donor tissues have a myenteric plexus with normal characteristics and typical morphology of ganglion cells (*data not shown*). While type II achalasia tissues and type II achalasia post-COVID-19 tissues had abundant and heterogeneous inflammatory infiltrates predominantly enriched with lymphocytes, scant polymorphonuclear leukocytes (plexitis), and scarce ganglion cells (Fig. 1 G).

### Tissue Proinflammatory Cells and Cytokines

IL-22 plays an essential role in host immunity and the maintenance of barrier function through the induction of innate antimicrobial at mucosal surfaces (epithelial immunity). Moreover, IL-22 synthesized by Th22 and/or Th17 cells contributes to autoimmune disease. The IL-17 is produced by Th17 cells, CD8 T cells, γδ T cells, neutrophils, macrophages, and various epithelial and parenchymal cells. IL-17A is a key mediator of autoimmune diseases. Under physiological and pathological conditions, Th17 cells induce B cell proliferation and differentiation into immunoglobulin-secreting cells and produce a wide range of pro-inflammatory cytokines and chemokines. It endorses the diapedesis of neutrophils and monocytes. The IFN-γ promotes cell-mediated immunity and activates mononuclear cells. Produced by activated Th1 cells and natural killer cells (NKs), IFN-γ regulates various immune and inflammatory responses. Specifically, this cytokine promotes immunomodulation and has anti-cancer and antimicrobial/antiviral activities. It inhibits type I collagen synthesis and produces chemokines and their receptors. The IL-22^+^ cell percentage was higher in the esophageal tissue of patients with type II achalasia regardless of whether they had been previously infected with the SARS-CoV-2 compared to transplant donors. The highest IL-22^+^ cell percentage was determined in patients with achalasia (Fig. 2A and B; Table 1). The Th17 (IL-17A^+^/CD4^+^; Fig. 2C and D; Table 1) and Th1 (IFN-g^γ^/CD4^+^; Fig. 3A and B; Table 1) cell percentage was similar in the LES muscle of patients with type II achalasia and those with COVID-19 before LHM, and it was higher and statistically significant compared with transplant donors.

**Figure 2.**
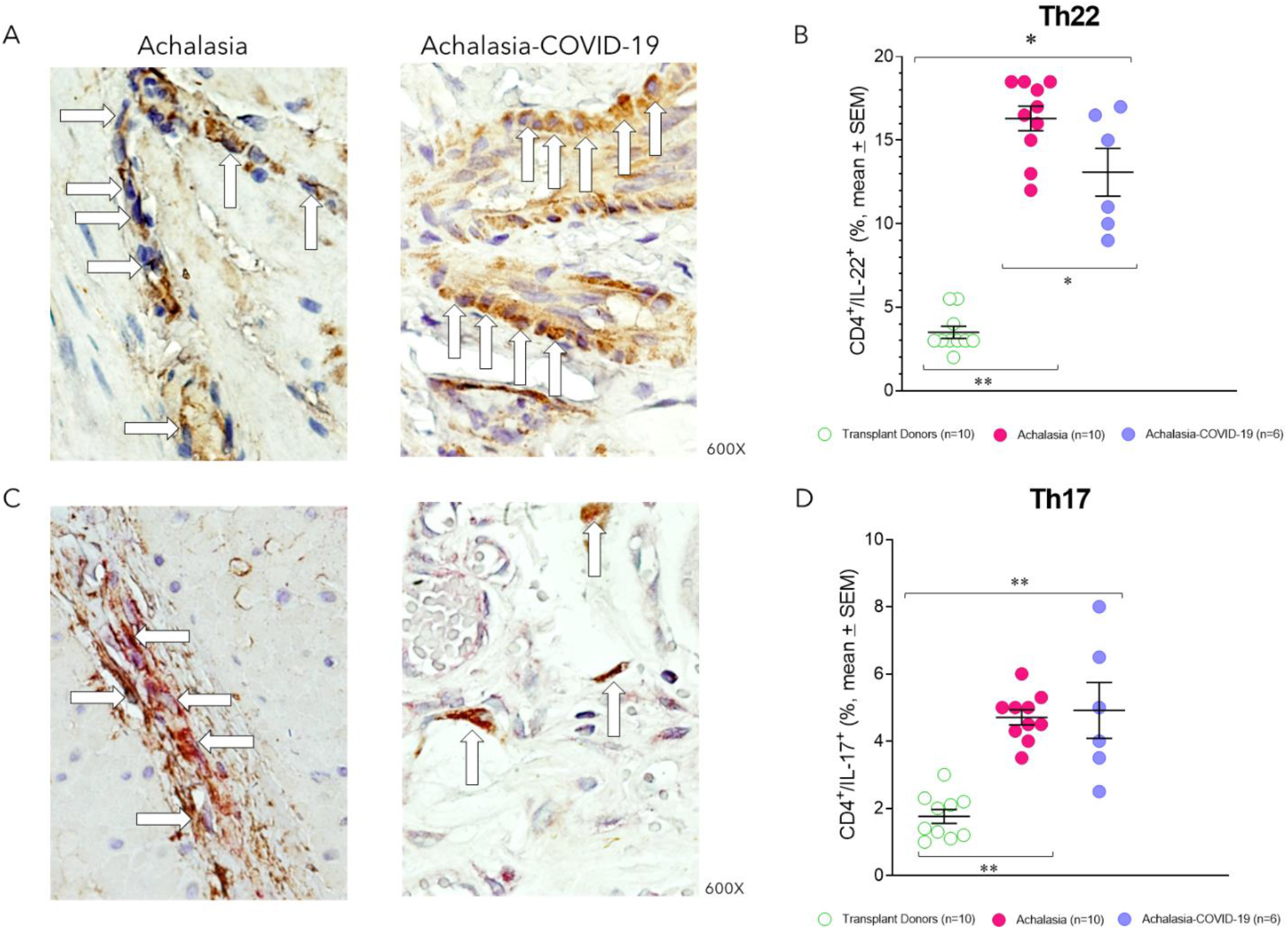
(A) Representative immunostaining of IL-22-expressing cells (brown) or (C) CD4+/IL-17-A+ double-positive cells (burgundy) in the LES muscle of type II achalasia (*left*) and type II achalasia post-COVID-19 patients (*right*). Arrows depict immunoreactive cells. The original magnification was x600. (B) Percentage of IL-22-expressing cells or (D) Th17 cells per microscopic field in the LES muscle of transplant donors (n=10), achalasia patients without previous infection of SARS-CoV-2 (n=10), and achalasia patients with the infection earlier of SARS-COV-2 (n=6). The results are expressed as mean ± SEM of positive immunoreactive cells. \**P*≤0.05; \*\**P*≤0.001.

**Figure 3.**
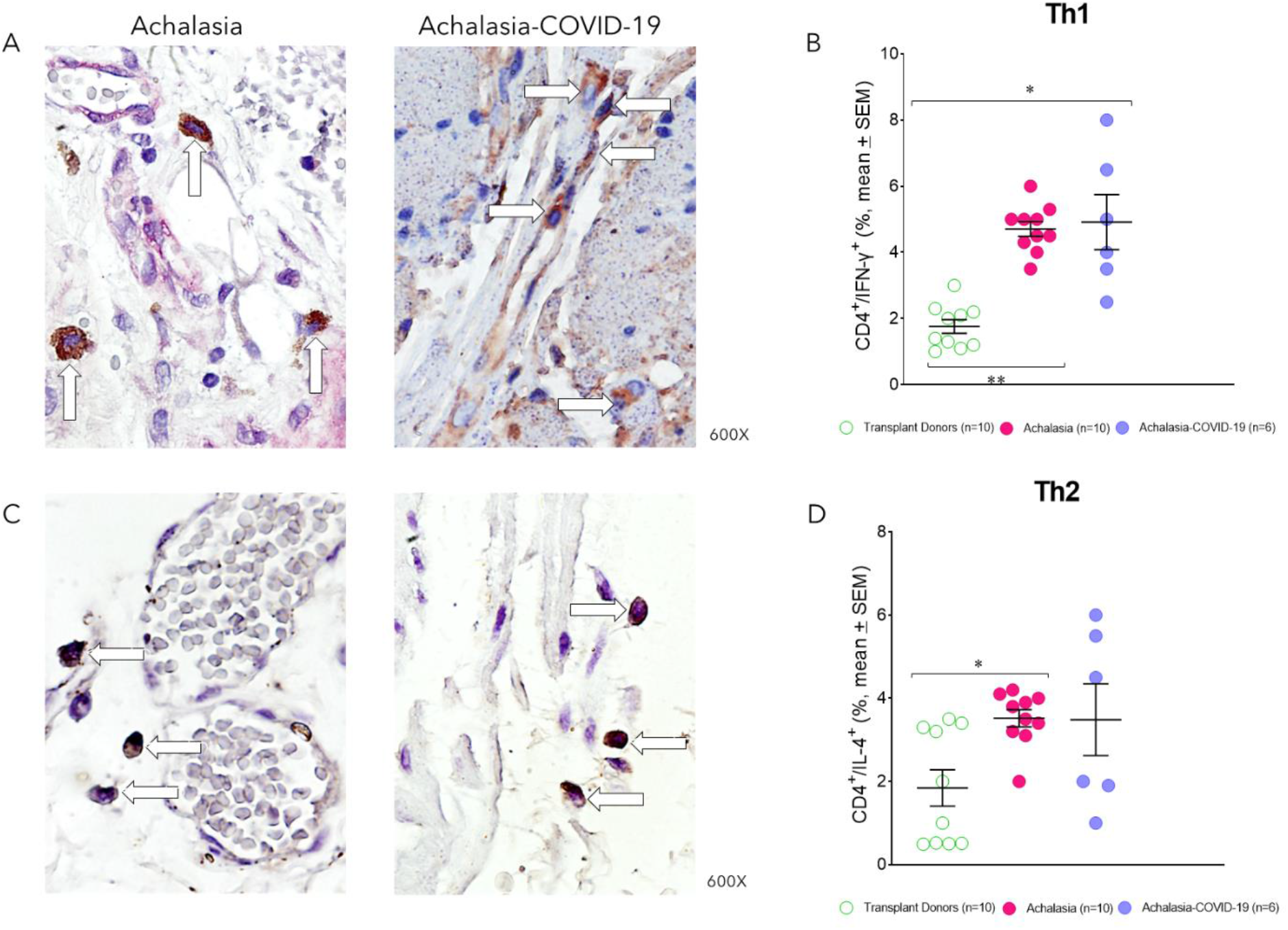
(A) Representative immunostaining of CD4/IFN-g-expressing cells (burgundy) or (C) CD4^+^/IL-4^+^ double-positive cells (burgundy) in the LES muscle of type II achalasia (*left*) and type II achalasia post-COVID-19 patients (*right*). Arrows depict immunoreactive cells. The original magnification was x600. (B) Percentage of Th1 cells or (D) Th2 cells per microscopic field in the LES muscle of transplant donors (n=10), achalasia patients without previous infection of SARS-CoV-2 (n=10), and achalasia patients with the infection earlier of SARS-COV-2 (n=6). The results are expressed as mean ± SEM of positive immunoreactive cells. \**P*≤0.05; \*\**P*≤0.001.

### Profibrogenic Tissue Cells

IL-4 is a fundamental cytokine during the fibrosis process. In addition, it abrogates the Th1 cell-and Th17 cell-mediated inflammation and differentiation and promotes Th2 cell differentiation. IL-4 is synthesized primarily by Th2 cells. The highest Th2 (IL-4+/CD4+) cell percentage was determined in patients with achalasia compared with those with COVID-19 before LHM and transplant donors (Fig. 3C and D; Table 1).

### Tissue Regulatory cells and Cytokines

CD4 regulatory T cells (Tregs) and IL-10-producing regulatory B-cells (Bregs) are specialized subpopulations of T and B cells that suppress the activation of CD4 and CD8 effector T cells. They maintain homeostasis and tolerance to autoantigens. The patients with type II achalasia who had COVID-19 before LHM had similar Treg (forkhead box P3-expressing CD25 T cells Fig. 4A and B; Table 1) and Breg (IL-10-expressing B regulatory cells Fig. 4C and D; Table 1) cell frequency with transplant donors, and lower cell number than achalasia patients (Fig. 4; Table 1).

**Figure 4.**
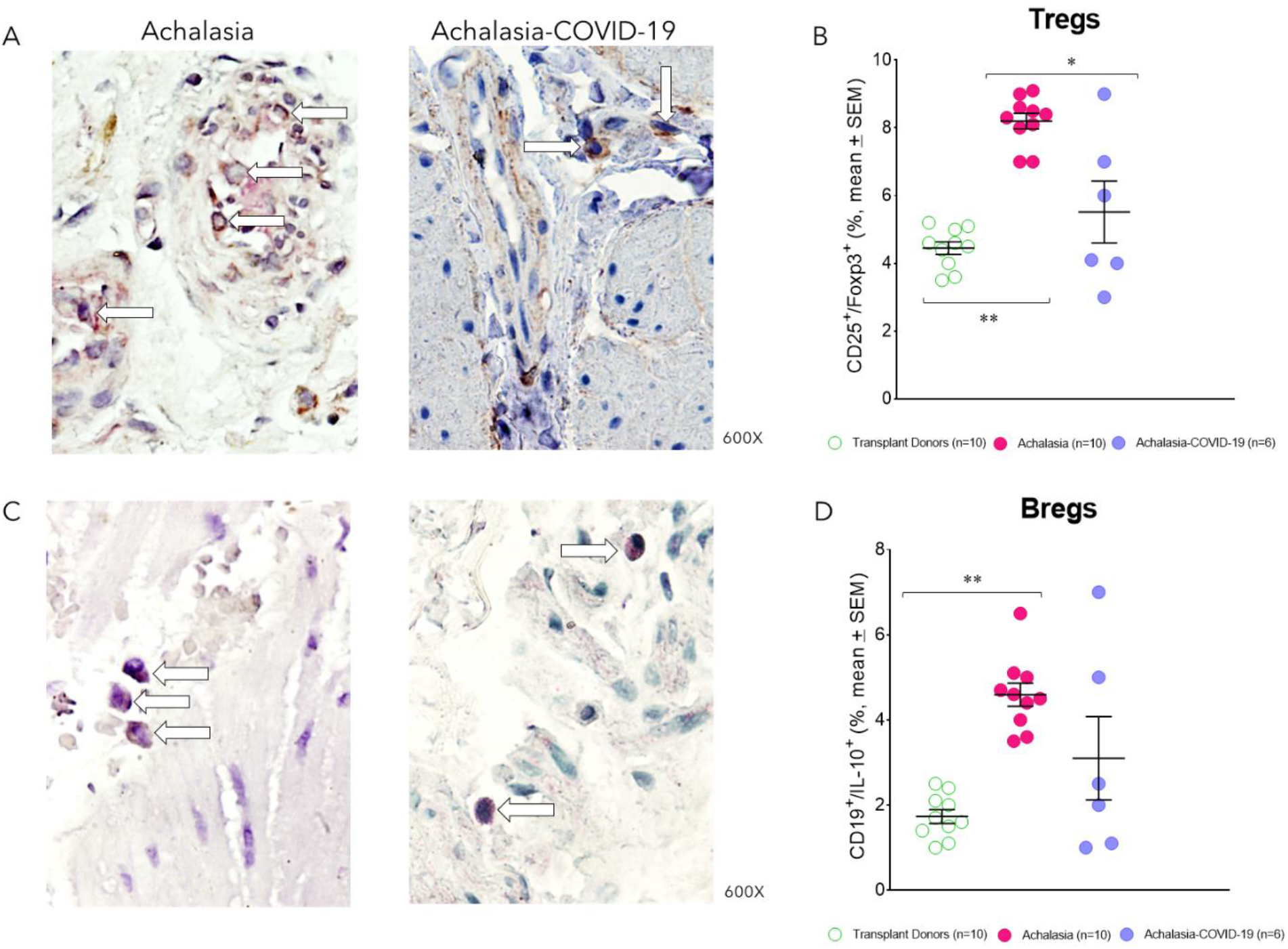
(A) Representative immunostaining of CD25/Foxp3-expressing cells (burgundy) or (C) CD20^+^/IL-10^+^ double-positive cells (burgundy) in the LES muscle of type II achalasia (*left*) and type II achalasia post-COVID-19 patients (*right*). Arrows depict immunoreactive cells. The original magnification was x600. (B) Percentage of Treg cells or (D) Breg cells per microscopic field in the LES muscle of transplant donors (n=10), achalasia patients without previous infection of SARS-CoV-2 (n=10), and achalasia patients with the infection earlier of SARS-COV-2 (n=6). The results are expressed as mean ± SEM of positive immunoreactive cells. \**P*≤0.05; \*\**P*≤0.001.

Plasmacytoid dendritic regulatory cells (pDCreg) are a subpopulation expressing indoleamine 2,3-dioxygenase (IDO) and inhibit T cell polarization and the differentiation of Tregs. The pDCreg cell percentage was similar in the LES muscle patients with type II achalasia and those with COVID-19 before LHM. It was higher and statistically significant compared with transplant donors (Fig. 5A and B; Table 1).

**Figure 5.**
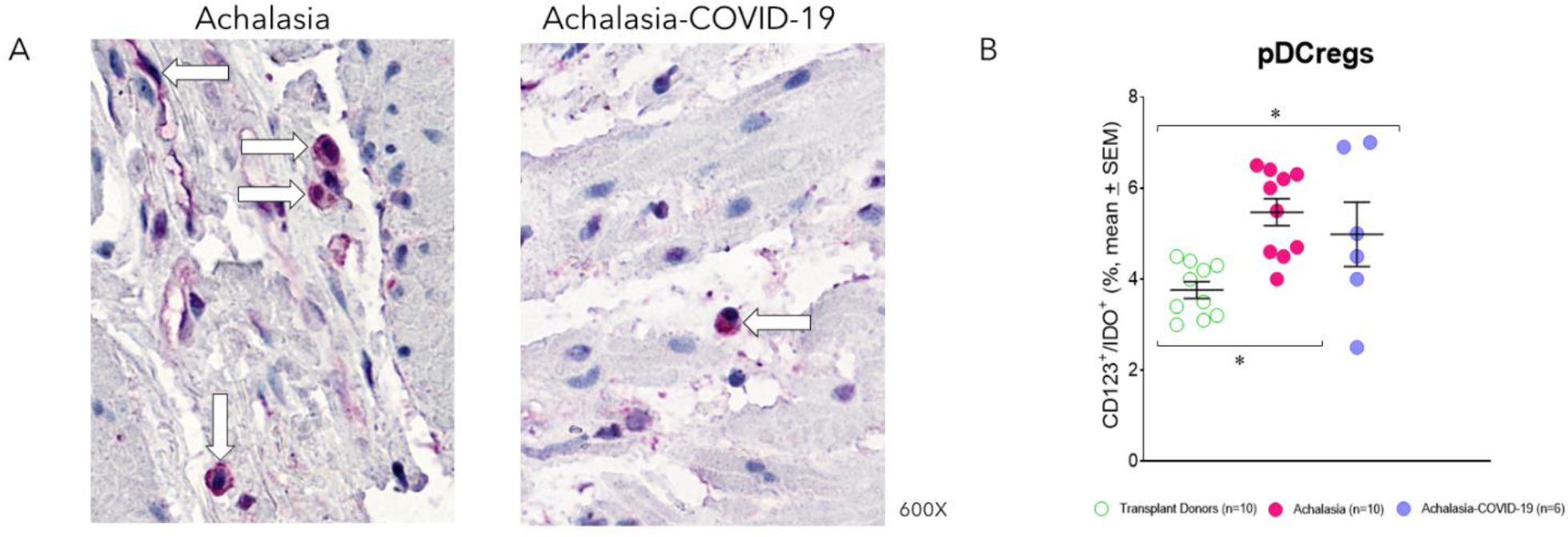
(A) Representative immunostaining of CD123/IDO-expressing cells (burgundy) in the LES muscle of type II achalasia (*left*) and type II achalasia post-COVID-19 patients (*right*). Arrows depict immunoreactive cells. The original magnification was x600. (B) Percentage of regulatory plasmacytoid dendritic cells per microscopic field in the LES muscle of transplant donors (n=10), achalasia patients without previous infection of SARS-CoV-2 (n=10), and achalasia patients with the infection earlier of SARS-COV-2 (n=6). The results are expressed as mean ± SEM of positive immunoreactive cells. \**P*≤0.05.

## Discussion

Increasing evidence indicates that SARS-CoV-2 infection is associated with neurological symptoms in a subgroup of patients with mild to severe COVID-19.^14,15^ Moreover, Esposito et al.^16^ have suggested that the enteric nervous system (ENS) could act as an entry route of SARS-CoV-2 to the brain. Thus, the virus would gain access to the brain via vagal and/or splanchnic nerves. A comparable mechanism of neurogenic transmission to the central nervous system was previously shown for herpes^17^ and influenza viruses.^18^ Interestingly, our group has detected *in situ* viral infection and active replication of HSV-1 in the LES muscle of patients with achalasia.^6^ The anatomical plausibility for SARS-CoV-2 neuro-invasion through the ENS has been demonstrated in a recent histochemical study on small and large intestinal specimens, choroid plexus, and adjacent brain parenchyma obtained post-mortem in COVID-19 patients.^19,20^ Moreover, ACE2 and TMPRSS2, two proteinases crucial for the entry of SASRS-CoV-2 into host cells, are abundantly expressed in the perikarya of enteric neurons and glial cells, both in the myenteric and submucous plexus of the small and large intestine, as well as epithelial cells. Enteric neurons show different levels of ACE2 staining intensity, suggesting a differential expression between neuronal subtypes.^20^

The disorders of gastrointestinal motor function up to severe motility derangement and pseudo-obstruction have been reported in half of the critically-ill COVID-19 patients with a high degree of systemic and intestinal inflammation.^21^

Additionally, a retrospective study evaluated 95 cases of COVID-19 patients for gastrointestinal complaints. It analyzed the virus’ presence by real-time RT-PCR assay in esophageal, gastric, duodenal, and rectal mucosal samples obtained endoscopically and fecal samples. This group found that 61% of the included patients presented gastrointestinal symptoms. Among the most frequent were diarrhea (24.2%), nausea (17.9%), anorexia (17.9%), acid reflux (2.1%), epigastric discomfort (2.1%), and upper gastrointestinal bleeding (2.1%). A positive correlation was found between severe and critical COVID-19 cases (defined as oxygen saturation ≤ 93% and a PaO2/FiO2 ≤ 300 mmHg) and the presence of SARS-CoV-2 RNA in all the gastrointestinal tract portions evaluated.^10^ The susceptibility of cells in the gastrointestinal tract to infection with SARS-CoV-2 provides a possible explanation for the presence of gastrointestinal symptoms in patients with acute COVID-19.

This study demonstrates the plausibility of infection of esophageal cells by SARS-CoV2 since we detect both the virus by PCR and immunohistochemistry (spike protein) and its ACE2 receptor. While the ACE2 receptor is expressed with higher or lower intensity in all LES tissues of patients with achalasia, regardless of whether the tissues belong to the pre-COVID-19 period or if biopsies were obtained from patients with the previous infection; the virus was detected in 6 of 7 patients who had COVID-19 diagnosed by PCR at least one month before surgery.

Concerning the profile of effector CD4+ T cells, it is very similar between the patients with achalasia type II who had COVID-19 before LHM and the group of patients with achalasia type II.

The most striking finding is that the number of regulatory T and B cells in the LES muscle patients with achalasia who had a previous infection by SARS-COV-2 was lower than in patients with type II achalasia like transplant donors. This suggests that inflammation’s mechanisms could surpass those of regulation in these patients, which could be associated with more severe disease.

Moreover, the case of a patient diagnosed with SARS-CoV-2 pneumonia, hospitalized, and that developed type II achalasia three months after moderate COVID-19 infection is striking. With all the limitations presented in this study, the fact that the patient developed the disease after COVID-19 infection and that HSV-1, SARS-CoV-2, and its receptor were detected in the LES muscle could suggest a link between viral infections and the development of achalasia. Viruses are considered critical biological agents that cause autoimmunity with mechanisms such as molecular mimicry, bystander activation of T cells, transient immunosuppression, and inflammation, which has also been seen in post-COVID-19 autoimmunity.^22,23^ In a previous study, our group has shown that most patients with achalasia have a latent and, in some cases, even active HSV-1 infection in the LES.^6^ Additionally, Facco et al. have identified an oligoclonal lymphocytic infiltrate within the LES of achalasia patients with the ability to recognize human HSV-1 antigens associated with an increased proliferation and Th1 type cytokines release.^24^ Some studies have demonstrated that a high incidence of herpesvirus reactivation is associated in patients admitted to the intensive care unit for severe COVID-19.^25,26^ Thus, according to de evidence, it is not preposterous to hypothesize that the SARS-CoV-2 virus might be a biological trigger agent for this autoimmune condition.^22^ However, long-term studies will shed light on this hypothesis to determine the possible etiopathogenic role of SARS-CoV-2 as a trigger agent for achalasia.

Hitherto there are no reports in the literature of achalasia as a complication of COVID-19. Even though this is a descriptive study, our findings are of interest because they may contribute to the knowledge about achalasia’s etiology and how the presence of the SARS-CoV-2 virus in the LES may be associated with the development of post-COVID-19 achalasia.

## Data Availability

All data produced in the present study are available upon reasonable request to the authors.

## GUARANTOR OF THE ARTICLE

Gonzalo Torres-Villalobos, MD, PhD.

## SPECIFIC AUTHOR CONTRIBUTIONS

<u>J.F.-C</u>., R.A.-S., A.TA, M.C.N.-P., E.B., E.C.-A., Conceptualization, Data curation, Formal analysis, Investigation, Methodology, Writing – original draft, Writing – review & editing, D.A.L., N.U.U., M.A.V., Conceptualization, Data curation, Formal analysis, Investigation, Methodology, Writing – original draft, Writing – review & editing, I.-C.M.E., C.-C.L.A., Conceptualization, Data curation, Formal analysis, Methodology, Writing – original draft, Writing – review & editing, FL- and <u>G.T.-V</u>., Conceptualization, Data curation, Formal analysis Investigation, Methodology Supervision, Writing – original draft, Writing – review & editing.

